# Snapshot hyperspectral fundus imaging system using a microlens array

**DOI:** 10.64898/2025.12.10.25341996

**Authors:** Jongchan Park, Liang Gao

**Affiliations:** Department of Bioengineering, University of California, Los Angeles, California 90095, USA; California NanoSystems Institute, University of California, Los Angeles, California 90095, USA

## Abstract

Retinal hyperspectral imaging holds significant promise for early disease diagnosis by quantifying the spectral signatures of metabolic and hemodynamic biomarkers. However, conventional hyperspectral imaging systems typically require extensive scanning, leading to prolonged acquisition times and rendering images susceptible to motion artifacts caused by involuntary eye movements. To address this limitation, we present a snapshot hyperspectral fundus camera employing a microlens array. The system features a streamlined optical architecture and compatibility with a standard commercial fundus camera across various field-of-view (FOV) configurations (20°, 35°, and 50°). Furthermore, the system allows a tunable balance between spectral resolution versus light throughput, enabling adaptation to a wide range of applications.

## 1. Introduction

The human retina offers a unique optical window into the body’s vascular and neurological health[1]. While standard color fundus photography remains the clinical gold standard for retina imaging, it fundamentally lacks spectral specificity. This is a critical limitation, as the light spectrum remitted from biological tissue (reflection and fluorescence emission) carries quantitative diagnostic information. Consequently, hyperspectral imaging (HSI), which acquires both spatial and spectral information from a sample, has gained significant attention for retinal imaging[2,3]. Numerous efforts have been made to develop clinically compatible hyperspectral systems aimed at identifying spectral signatures associated with pathologies such as diabetic retinopathy[4], Alzheimer’s disease[5], and age-related macular degeneration[6].

Despite the diagnostic utility of these spectral biomarkers, integrating conventional hyperspectral imaging architectures into a clinical fundus camera presents significant challenges. Acquiring a three-dimensional hyperspectral datacube (*x,y,λ*) using conventional image sensors typically requires extensive scanning in either the spatial or spectral domain[7]. For instance, a hyperspectral line-scan camera disperses light using a prism or diffraction grating orthogonal to the line direction, measuring a single spatio-spectral slice (*x,λ*) per readout. Alternatively, a wavelength-scanning camera captures one spatial slice (*x,y*) at a fixed wavelength (*λ*) by using a tunable spectral filter and scans across the entire spectral range. However, these sequential acquisition requirements result in prolonged scan times, which are highly unfavorable for *in vivo* retinal imaging. The human eye exhibits constant involuntary motion, including tremors, drifts, and microsaccades, with timescales ranging from tens to hundreds of milliseconds. Consequently, traditional scanning-based techniques frequently suffer from motion artifacts, which can confound spatial and spectral signatures and render quantitative analysis inaccurate.

A promising solution for hyperspectral retinal imaging is the use of snapshot hyperspectral imagers, which capture 3D datacubes in parallel, making them immune to motion blur during exposure[8–10]. One prominent example, the Image Mapping Spectrometer (IMS), captures the snapshot scene by employing a custom-designed mirror array, known as an image mapper[11,12]. This component consists of multiple angled mirror facets that redirect different parts of an image to distinct regions on a detector array. This redirection creates void spaces between image slices, allowing a prism or diffraction grating to spectrally disperse light orthogonal to the length of each slice without overlap. Consequently, the IMS captures a spectrum from every spatial location within a single exposure, and the original image is reconstructed through straightforward pixel remapping. However, imaging performance is strictly limited by the fabrication precision of the custom image mapper. Issues such as striping artifacts arising from non-uniform mirror reflectivity, ghost images caused by crosstalk between spectral channels, and the high complexity of fabrication and calibration have hindered its widespread adoption[13].

Various computational imaging techniques have been developed for hyperspectral fundus imaging which have distinct pros and cons over IMS. For example, the Computed Tomographic Imaging Spectrometer (CTIS) has been demonstrated and integrated into standard fundus camera setups[14]. These systems use a computer-generated holographic element to duplicate and disperse the scene into multiple projections. The hyperspectral scene is then reconstructed by solving the associated inverse problem. However, the limited number of projections and the “missing cone” problem in tomography often lead to blur and artifacts. The Coded Aperture Snapshot Spectral Imager (CASSI) is another promising method, which encodes the hyperspectral scene with a coded mask prior to dispersion[15,16]. The detector captures a spatially and spectrally overlapped scene, and the hyperspectral datacube is computationally reconstructed. Although CASSI can capture datacubes with voxel counts exceeding the detector’s pixel count, the ill-posed nature of the image formation model makes the system susceptible to noise and reconstruction artifacts[17,18].

In this work, we propose a snapshot hyperspectral fundus imaging system based on the principle of integral field spectroscopy using a microlens array (MLA) [8,19,20]. The system can capture a hyperspectral datacube of size 240×240×31 (*x, y, λ*) in a single exposure. The observed spectrum ranges from 450 nm to 750 nm, providing an average spectral sampling rate of 9.7 nm per pixel. The system features a streamlined optical architecture compatible with commercial fundus cameras and supports various FOV configurations. Furthermore, a variable slit mechanism enables a tunable balance between light throughput and spectral resolution, allowing the measurement scheme to be optimized for specific clinical applications.

## 2. Methods

### 2.1 Hyperspectral imaging principle

The operating principle of our hyperspectral imaging system is based on integral field spectroscopy, a technique widely employed in astronomy[19]. In this method, the incident scene is spatially sampled and mapped onto distinct spatial locations using an integral field unit, such as image slicing mirrors[11], fiber arrays[21], or a lenslet array[22]. The reformatted image is then spectrally dispersed onto a detector array in a manner that avoids spatial-spectral overlap.

Among the various integral field unit types, we utilized an MLA for this implementation. Although snapshot hyperspectral imagers based on image slicers have been demonstrated in retinal imaging[23], the fabrication of high-precision micro-mirror facets for the optical regime has hindered widespread adoption. These mirrors typically feature high aspect ratios, with widths of tens of micrometers and heights of several tens of millimeters, requiring tilt angles tailored with high precision. Furthermore, striping artifacts caused by non-uniform reflectivity of the mirror facets are detrimental to clinical applications. While post-processing techniques can mitigate these artifacts, they often compromise quantitative accuracy, particularly under the photon-starved conditions typical of fundus imaging.

In contrast, high-quality MLAs are now readily available in both off-the-shelf and custom configurations, driven by recent advancements in photolithography and 3D printing[24]. These arrays are typically fabricated on flat glass substrates and are easily integrated into optical systems. In our system design, the MLA is positioned at the focal plane of the imaging lens. It generates a series of pupil images, where each spot corresponds to the light integrated over the spatial aperture of a single lenslet. To ensure a compact form factor while preserving high light throughput, a large-format detector is positioned directly behind the low-f-number MLA (Fig. 1).

**Fig. 1.**
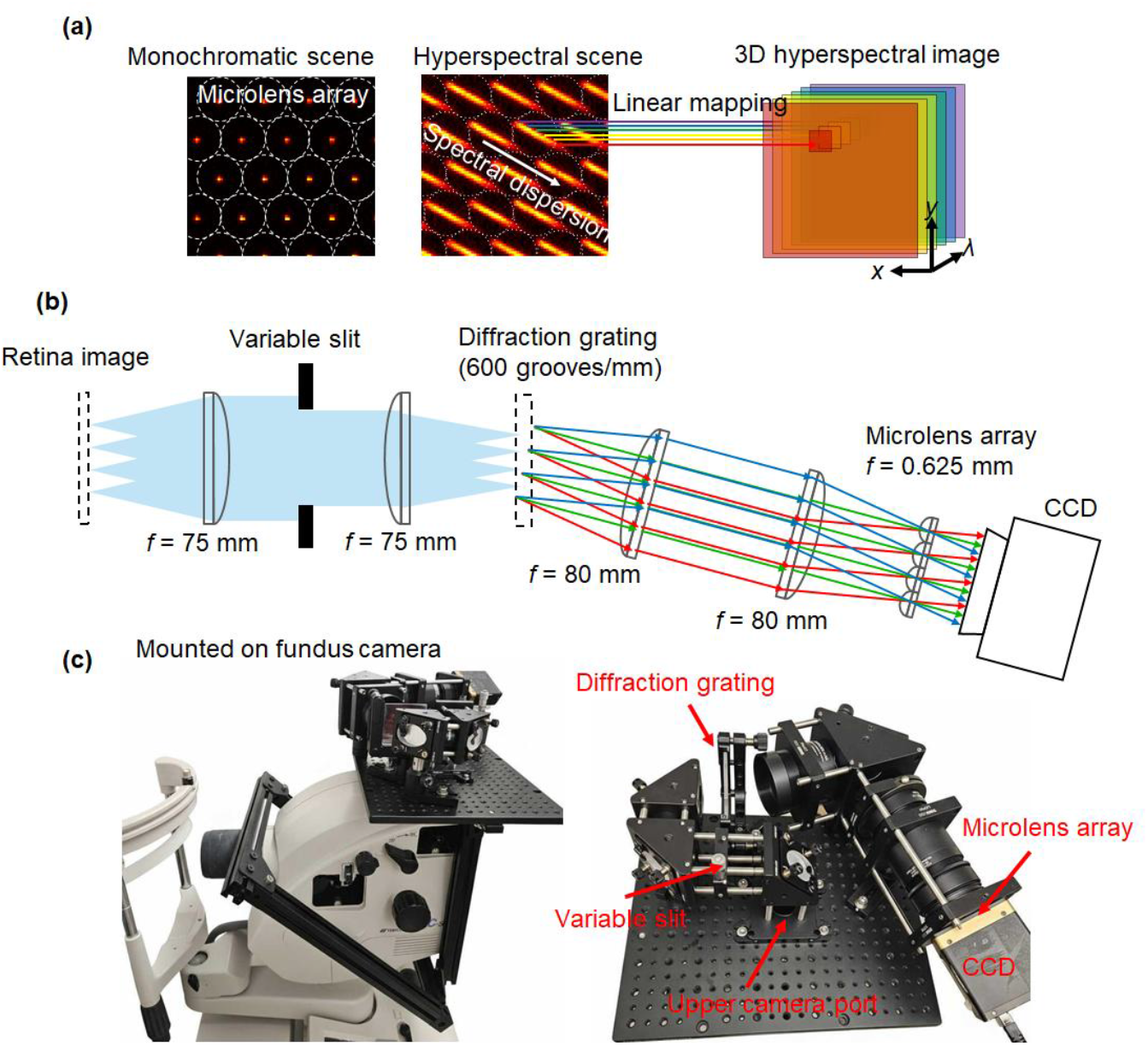
Hyperspectral fundus imaging system. (a) Hyperspectral imaging principle. The MLA spatially samples the scene, and the grating disperses the scene into non-overlapping spectral directions on the sensor. The hyperspectral scene is reconstructed by a linear mapping. (b) Optical layout of the system. (c) Photogram of the hyperspectral fundus imaging system. The system is mounted on the upper imaging port of a commercial fundus camera (Topcon, TRC-50DX).

### 2.2. Optical setup

The hyperspectral imaging module was designed as an add-on attachment to the upper imaging port of a standard fundus camera (TRC-50DX, Topcon Healthcare). The optical schematic is illustrated in Fig. 1(b). The internal halogen lamp of the fundus camera illuminates the retina with an annular pattern, which suppresses the stray light caused by the cornea and provides uniform illumination over the retina.

The reflected light from the retina passes through a spectral filter (FESH0750, shortpass filter, cut-off wavelength 750 nm, Thorlabs) and forms an image at the upper imaging port. Two achromatic lenses (AC254-075-A, Thorlabs) relay the image to the transmissive diffraction grating (GT50-06V, Thorlabs). A variable slit (VA100CP, Thorlabs) is located at the Fourier plane, where the slit direction is orthogonal to the dispersion direction of the diffraction grating. The dispersed image is relayed by another pair of achromatic lenses (AC508-080-A, Thorlabs). An MLA (focal length, f = 625 µm, diameter, d = 100 µm) is placed at the focal plane of the imaging lens. The scene is captured by a high-resolution CCD camera (B6640, Imperx, 6600 × 4400 pixel resolution, 5.5 µm pixel pitch).

As illustrated in Fig. 1(b), the MLA spatially samples the scene from the fundus camera into discrete spots. The microlenses are arranged in a honeycomb pattern. The diffraction grating disperses the spots along a 30-degree tilt angle. This configuration maximizes the utilization of the spatial pixels in the CCD while ensuring that spectral information from adjacent spatial points does not overlap on the camera plane. The 3D hyperspectral datacube is captured in a single snapshot measurement and the reconstruction is performed via linear operations. The prototype of the system integrated with the fundus camera is shown in Fig. 1(c). In order to ensure that the hyperspectral imaging system does not interfere with the standard operation of the fundus camera, the system is built with a modular design and mounted on the upper camera port of the commercial fundus camera.

### 2.3 System calibration

We calibrated the spectral response of the imaging system by using a series of spectral bandpass filters ranging from 450 nm to 700 nm (Fig. 2(a)). A second-order polynomial fit was applied to find the relation between the spectral channel, which is the pixel displacement along the dispersion axis, and the actual wavelength. Approximately 30 spectral channels capture the spectrum in the range of 400 nm to 750 nm. We note that the spectral dispersion curve is non-linear and the dispersion corresponding to a single camera pixel at 400 nm and 750 nm is 4.5 nm and 18 nm, respectively. The spectral resolution of the system is determined by the diameter of the microlens, camera pixel pitch, and dispersion power of the diffraction grating. In our system configuration, the spectral resolution is limited by the digital sampling rate.

**Fig. 2.**
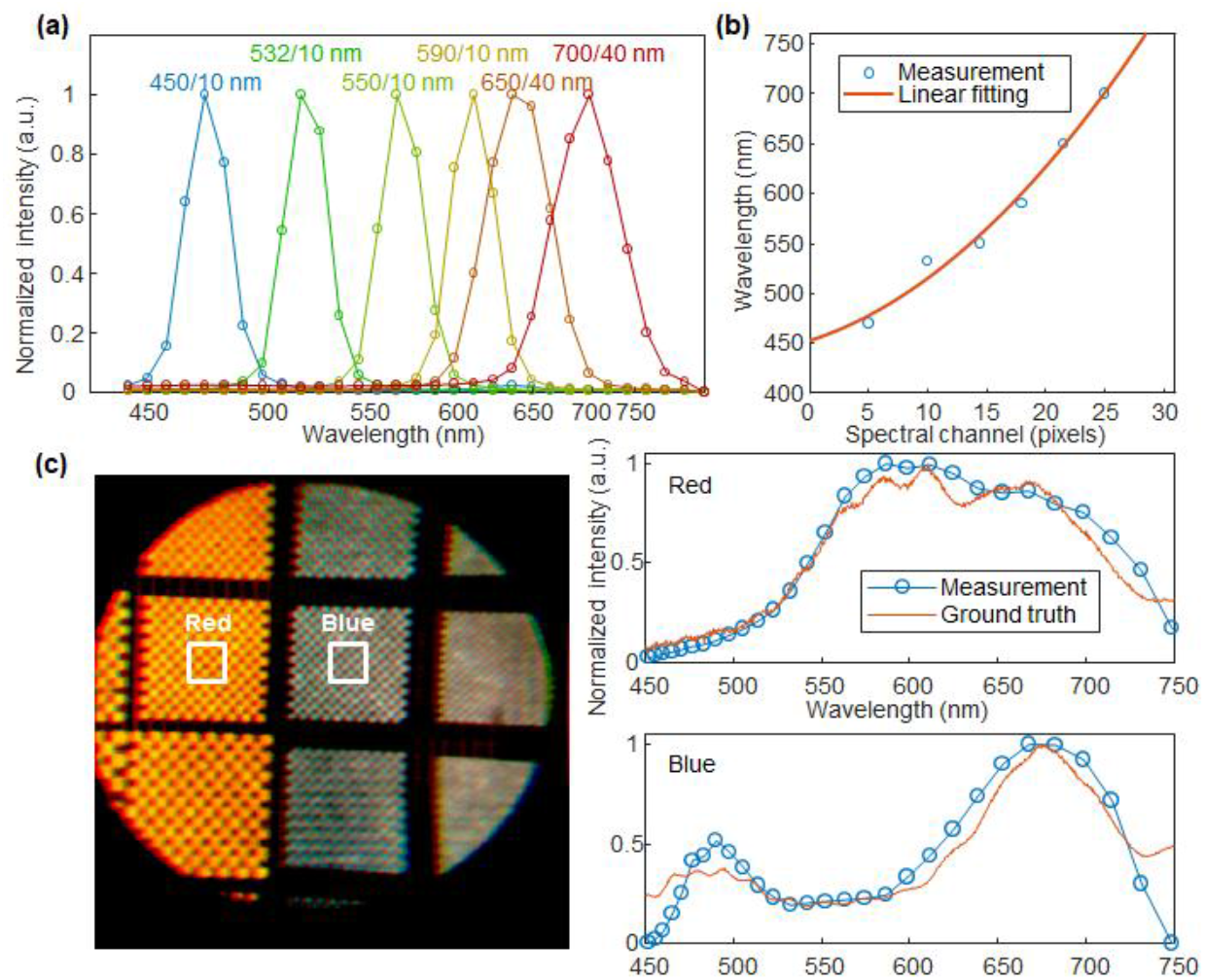
Spectral calibration and validation. (a) Spectral response calibration using bandpass filters. (b) Calibration curve showing the polynomial relationship between spectral channel (pixel displacement) and wavelength. (c) Validation against a color checker. Left: Reconstructed false-color image. Right: Comparison of measured spectra (blue circles) vs. ground truth (orange line) for red and blue regions.

The spatial resolution of the system is determined by the number of microlenses that capture the scene. The image size at the upper port of the fundus camera is approximately 25 mm. The image is relayed with one-to-one magnification and projected onto the MLA of diameter 100 µm. The image is then captured by the camera with the format of 36.3 mm × 24.2 mm, which leads to a spatial sampling rate of approximately 250 × 242. Because of the limited clear aperture of relaying optical lenses and imperfections in the alignment, we extract the hyperspectral datacube with a spatial resolution of 240 × 240 throughout the experiment.

To validate the spectral accuracy, we imaged a color checker constructed using colored films (blue and red). Fig. 2(c) shows a false-color image reconstructed by using the measured spectrum in the range of 400 nm to 750 nm projected onto the CIE 1931 chart. The image appears to be more yellowish because the checker was illuminated by the halogen lamp. The spectral irradiance of the scene captured by our imaging system was compared to the ground truth captured by a benchmark fiber spectrometer (L-25-400-SMA, Ocean Optics). The measured spectra are consistent with the ground truth values. The spectral irradiance drops near the end of the spectrum (750 nm) where the shortpass filter edge is located.

## 3. Results and discussion

### 3.1 Eye phantom imaging

To validate the retinal imaging capabilities of the system, we imaged a standard eye phantom (Wide-field model eye, Rowe Technical design) (Fig. 3). It models an emmetropic eye with a 5 mm pupil diameter. The vascular-like patterns are printed on the retinal surface. We captured the scene in snapshot mode with 20°, 35°, and 50° FOV configurations. The illumination was provided by the fundus camera’s internal halogen lamp, which is a low-intensity light source conventionally used for viewing and alignment. The exposure time was set to 100 ms. The captured 2D image was processed to reconstruct the hyperspectral scene with a size of 240×240×31 (*x, y, λ*).

**Fig. 3.**
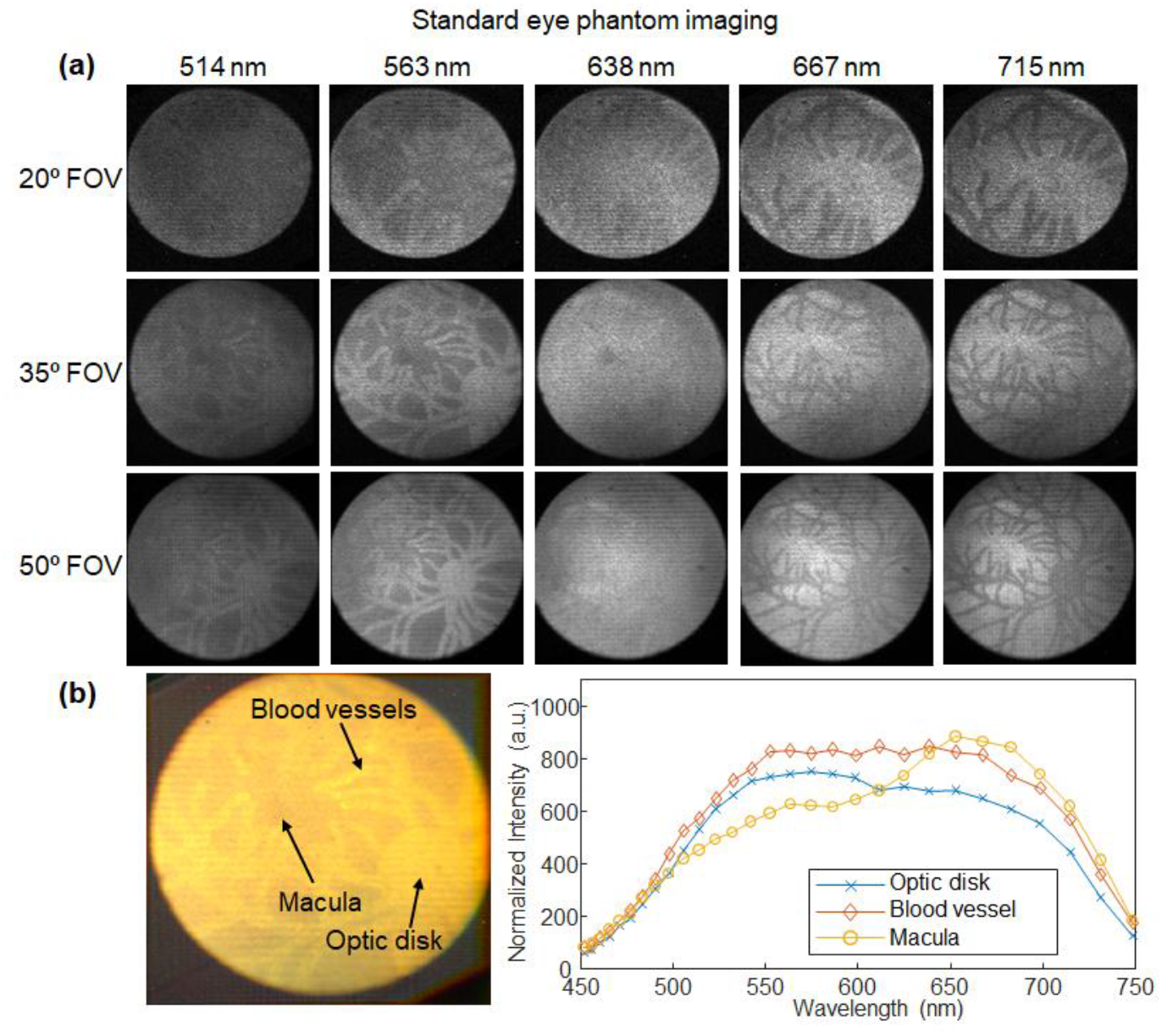
Standard eye phantom imaging. (a) Snapshot hyperspectral images of the retina across three fields of view (20°, 35°, 50°). Columns show reconstructed monochromatic images at selected wavelengths (514–715 nm). (b) Spectral analysis. Left: False-colored image from 35° FOV measurement. Right: Mean spectral intensity profiles for the optic disk, blood vessels, and macula mimicked regions.

Figure 3(a) illustrates the representative monochromatic spectral channels reconstructed from the single snapshot measurements. The images demonstrate high spatial fidelity and clearly resolve fine anatomical structures. The different retinal structures exhibit varying reflectance across wavelengths. For instance, retinal vessels are highly reflective in the green/yellow bands (563 nm) while they appear to be dark in the red/NIR wavelengths (715 nm), revealing underlying structures.

We extracted average spectral signatures from specific regions of interest (blood vessels, macula, and optic disk mimicked areas) as indicated in the false-colored image shown in Fig. 3(b). Although the standard eye phantom is different from the *in vivo* model, we can still identify distinct spectral signatures in the measurements. The macula shows reduced blue/green reflectance mimicking the macular pigment absorption, and blood vessels show a relatively flat spectrum. The optic disk, known to have high reflectance, is not represented in this eye phantom model.

### 3.2 Discussion

Optical light throughput is a critical factor in a hyperspectral fundus camera because the system requires capturing a large amount of spectral data, which often leads to a low signal-to-noise ratio. In addition, the light dose should be limited to avoid potential photodamage to the retina. In our system, we can capture the hyperspectral scene with maximum light throughput by fully opening the variable slit. However, this increases the image size at the Fourier plane and results in compromised spectral resolution.

We evaluated the impact of slit width on spectral resolution using monochromatic 532 nm illumination across different FOVs (20°, 35°, and 50°). Figure 4(a) compares the raw sensor output with the slit fully open and with a 1 mm width. We note that the image at the Fourier plane is non-uniform, which differs from conventional cameras. This is attributed to the annular illumination unit and the alignment aperture located in the beam path of the commercial fundus camera. When the slit is fully open, the image size at the Fourier plane is approximately 1.5 mm, 2.6 mm, and 3.8 mm at the 20°, 35°, and 50° FOV configurations, respectively. This results in decreased spectral resolution, as these image sizes exceed the effective single-pixel pitch of the camera, 0.66 mm at the slit plane. Conversely, when the slit is closed to 1 mm, the spectral resolution is limited primarily by the spatial sampling of the camera.

**Fig. 4.**
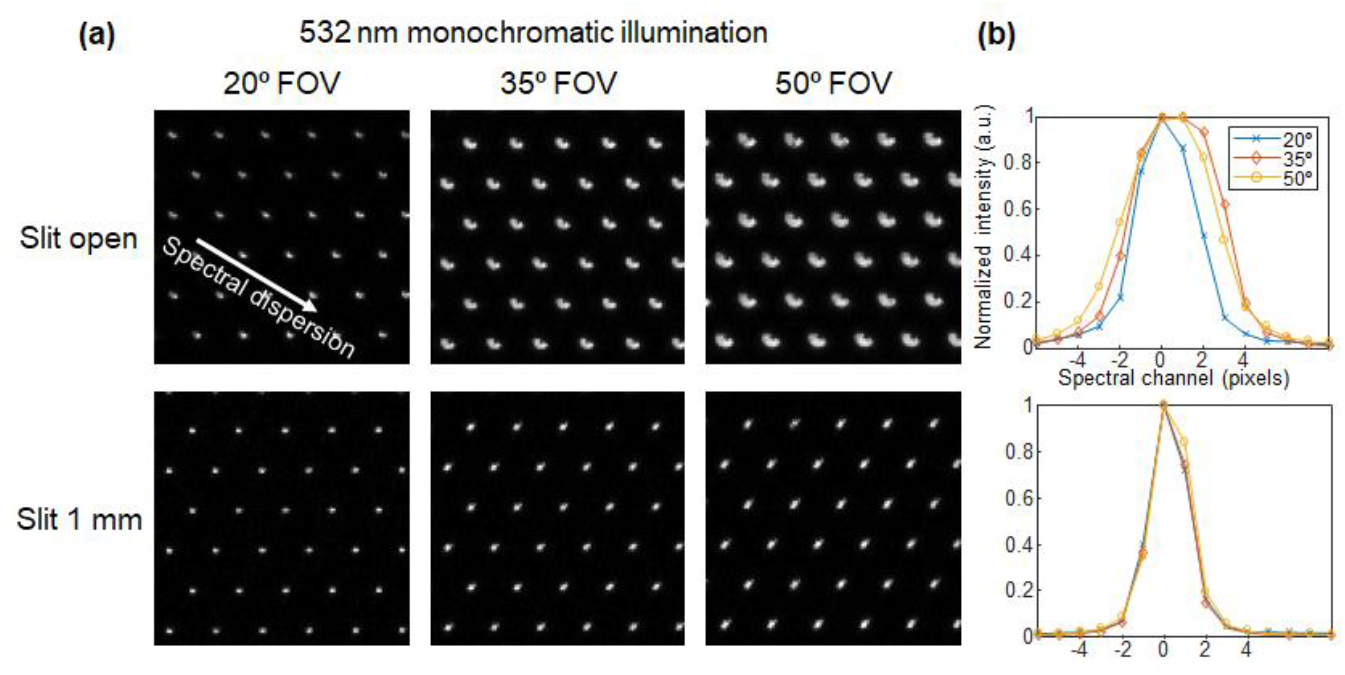
System characterization at varying slit width. (a) Raw sensor images of 532 nm monochromatic illumination at 20°, 35°, and 50° FOVs. Comparing “Slit open” (top row) vs. “Slit 1 mm” (bottom row) shows a significant reduction in spot size with the narrower slit. (b) Normalized intensity profiles of the spectral point spread function (PSF) for the slit-open (top) and slit-1mm (bottom) configurations.

Notably, for the 20° FOV, high light throughput is maintained even when the slit is narrowed to 1 mm. Even for larger FOV configurations, the use of a slit enables signal integration along the axis orthogonal to the spectral dispersion direction, making the system more photon-efficient than using a pinhole aperture[25]. This tunability allows the operator to prioritize either spectral resolution (narrow slit) or signal-to-noise ratio (open slit), depending on the specific clinical application.

## 4. Conclusion

We have demonstrated a snapshot hyperspectral fundus imaging system integrated with a commercial clinical fundus camera. By utilizing an MLA and diffraction grating, the system captures hyperspectral datacubes with high fidelity in a single exposure, thereby mitigating motion artifacts. The integration of a variable slit provides tunable control over light throughput and spectral resolution. Imaging results using a standard eye phantom confirm the system’s capability to resolve spectral biomarkers even under low-intensity halogen lamp illumination. Given its high light throughput and compatibility with commercial fundus cameras, the system holds great promise for clinical translation.

## Data Availability

All data produced in the present study are available upon reasonable request to the authors

## Funding

National Institutes of Health (R35GM128761, R01HL16531, R01NS142690); Imaging Research in Alzheimer’s and other Neurodegenerative diseases from The Alzheimer’s Association & The Foundation of the American Society of Neuroradiology (ASNR).

## Data availability

Data underlying the results presented in this paper are not publicly available at this time but may be obtained from the authors upon reasonable request.

## Notes

### Competing Interest Statement

The authors have declared no competing interest.

### Funding Statement

National Institutes of Health (R35GM128761, R01HL16531, R01NS142690), Imaging Research in Alzheimers and other Neurodegenerative diseases from The Alzheimers Association & The Foundation of the American Society of Neuroradiology (ASNR).

